# Pathologic aggregation of α-synuclein and tau in the gut mirrors the brain, in both Lewy body disease and non-synucleinopathies

**DOI:** 10.64898/2026.11.19.26367659

**Authors:** Gregory D. Scott, Allison Quinn, Savanah Sicard, Anna Bui, Madison Christopher, Alaaeddin Alrohaibani, Peter Kay, Kristin Piché, Ariel Louwrier, Ursula S. Sandau, Randy Woltjer, Joseph F. Quinn

**Author notes:** Corresponding Author: Gregory Scott Pathology School of Medicine Administrative Office 3181 S.W. Sam Jackson Park Road, L-113 Portland, OR 97239. Co-first authors (Gregory D. Scott, Allison Quinn). **Relevant conflicts of interest/financial disclosures:** none. **Funding sources for the study:** This work was supported by VA Biomedical Laboratory Research and Development Career Development Award 1 IK2 BX005760-01A1, John and Tami Marick Family Foundation, Collins Medical Trust Award, Oregon Health & Science University Medical Research Foundation New Investigator Award, Parkinsons Center of Oregon Pilot Award, and VA military exposures research program (MERP) supplement to BLRD Merit I01 BX006155 (to GDS). The work was also supported by the Oregon Brain Bank and the Parkinson’s Disease and Mental Illness Research, Education, Clinical Center (PADRECC and MIRECC) at the Portland VA.

## Abstract

Seeded aggregation of α-synuclein and tau in the gastrointestinal tract is a potential biomarker and window into disease pathogenesis, but its quantitative relationship to the brain is unknown. Here, various regions of the gut and brain from autopsies of Lewy body disease and other diagnoses were tested for seeded aggregation of α-synuclein and an Alzheimer’s disease-related tau species, tau-dGAE. Gut seeding of α-synuclein (51.8 ± 16.8 vs 112.6 ± 23.0 hours) and tau-dGAE (28.5 ± 2.6 vs 31.7 ± 2.9 hours) was faster in Lewy body disease than non-Lewy cases and was correlated with seeding in frontal cortex and midbrain/medulla (Pearson’s r from 0.75 – 0.63). Seeding of α-synuclein in gut tissue was also correlated with loss of nigral pigment, pigmented neurons, Lewy bodies, and tyrosine hydroxylase-positive neurons as well as α-synuclein in the dorsal motor nucleus of the vagus (r values from 0.76 – 0.49).

Seeding of tau-dGAE in duodenum (but not brain) was faster in Lewy body disease compared to non-Lewy cases (28.5 ± 2.6 vs 31.7 ± 2.9 hours). Tau seeded aggregation in duodenum was correlated with tau-dGAE seeded aggregation in hippocampus and frontal cortex (r values from 0.58 - 0.55) and with tau and amyloid immunohistochemistry in the hippocampus, mainly for cases of Lewy body disease. Seeding for α-synuclein and tau-dGAE in the gut correlated with seeding in the brain regardless of disease status. Overall data show a quantitative mirroring of peripheral and central seeded aggregation and neuropathology and demonstrate its utility for studies of pathogenesis and clinical biomarkers.

## INTRODUCTION

Seeded aggregation of pathologic proteins offers a view into the prion-like pathogenesis of neurodegenerative disease and serves as a clinical biomarker. The gastrointestinal (GI) system is a useful peripheral target for studying seeding in Lewy body disease (LBD) by having early and prevalent symptoms, routine biopsy in the general healthy population, and alterations to the enteric nervous system that mirror central pathology. Prior research using seeding assays in peripheral body sites have typically tuned the method for binary classification. Consequently, there are functional knowledge gaps about how peripheral and central pathologic proteins relate to each other and to core neuropathologic features. For example, in terms of α-synuclein the quantitative relationship between peripheral (e.g. gut) and central seeding is unknown and it is unclear how it changes in the context of LBD and if it relates to central loss of dopaminergic neurons and Lewy pathology. Furthermore, the rostro-caudal gradient of α-synuclein abundance, where immunohistochemical α-synuclein burden is greater in the upper gastrointestinal tract (e.g. duodenum) versus lower tract (e.g. colon) has had limited testing in terms of functional seeded aggregation. Less is known about tau, which has not been measured in humans in terms of seeding in the gastrointestinal tract and neurodegenerative disease. Such data would help clarify if peripheral seeding of tau relates to Alzheimer’s Disease (AD) co-pathology in the context of LBD. Finally, it is unresolved if archived formalin-fixed and paraffin embedded tissues could be used to address these questions given their technical challenges with seeding assays. Overall, such data would help clarify the functional inter-dependencies of pathologic proteins throughout the body in neurodegenerative diseases.

To address these knowledge gaps, the current study includes subjects at autopsy with or without LBD and measures seeding of α-synuclein and AD-related tau (tau-dGAE) in archival GI tract and brain tissues. Data were collected at multiple GI sites and seeding was compared to diagnosis, other sites, and to histologic features of neurodegeneration and diagnostic neuropathology.

## METHODS

### Patient Samples

Tissues from forty-two participants were obtained sequentially at autopsy from The Oregon Brain Bank at Oregon Health and Science University (OHSU IRB No. 1623). Samples of duodenum, appendix, large intestine, midbrain/medulla, hippocampus, and frontal cortex were collected, fixed in formalin, and embedded in paraffin. All subjects underwent neuropathologic evaluation by a board-certified neuropathologist (RW). Twenty-three subjects received a neuropathologic diagnosis of Lewy Body Disease (LBD), and 19 control subjects did not (non-LBD). The LBD group included subjects with clinical diagnoses of Parkinson’s disease (PD), Dementia with Lewy Bodies (DLB), Lewy Body Disease, Not Otherwise Specified (LBD-NOS). The control group included subjects with Alzheimer’s Disease (AD), Hippocampal Sclerosis (HS), Progressive Supranuclear Palsy (PSP), mild ischemic injury (MII), or no diagnostic pathology (**Table 1**).

**Table 1:** Cohort characteristics and neuropathology.

|  |  |  |  |  |  | seeded amplification assay performed |  |  |  |  |
|---|---|---|---|---|---|---|---|---|---|---|
| No. | Group | Diagnosis | Age bin<br>(years) | Sex | Post-mortem<br>interval (hours) | Duodenum<br>aSyn & tau | Lrg intestine<br>aSyn | Fr. cortex<br>aSyn & tau | Midbr/Medulla<br>aSyn | Hippocamp<br>tau |
| 1 | lbd | ad pd lbd | 76-80 | f | 8 | x | x | x | x |  |
| 2 | lbd | ad pd lbd | 76-80 | f | 17 | x | x | x | x |  |
| 3 | lbd | ad pd lbd | 81-85 | f | 17 | x | x |  | x |  |
| 4 | lbd | ad lbd hs | 76-80 | m | 15 | x |  | x | x |  |
| 5 | lbd | ad lbd | 81-85 | m | 4 | x | x | x | x | x |
| 6 | lbd | ad lbd | 81-85 | f | 22 | x | x | x | x | x |
| 7 | lbd | ad lbd | 71-75 | m | 5 | x | x | x | x | x |
| 8 | lbd | ad lbd inf | 91-95 | f | 21 | x | x | x | x | x |
| 9 | lbd | lbd pd | 66-70 | m | 14 | x | x | x | x | x |
| 10 | lbd | pd lbd | 71-75 | m | 6 | x |  |  | x |  |
| 11 | lbd | pd lbd | 61-65 | f | 24 | x | x | x | x |  |
| 12 | lbd | pd lbd | 76-80 | m | 5 | x | x | x | x |  |
| 13 | lbd | lbd | 66-70 | m | 16 | x | x | x | x |  |
| 14 | lbd | lbd | 76-80 | f | 24 | x | x | x | x |  |
| 15 | lbd | lbd | 66-70 | m | 22 | x | x | x | x | x |
| 16 | lbd | lbd | 71-75 | m | 28 | x | x |  | x | x |
| 17 | lbd | lbd inf | 76-80 | m | 24 | x | x | x |  | x |
| 18 | lbd | lbd | 81-85 | m | 5 | x | x | x | x |  |
| 19 | lbd | pdd | 76-80 | m | 36 | x | x | x | x | x |
| 20 | lbd | pdd | 71-75 | m | 15 | x |  |  | x |  |
| 21 | lbd | pd | 71-75 | m | 33 | x | x | x | x | x |
| 22 | lbd | lbd | 81-85 | m | 27 | x | x | x | x |  |
| 23 | lbd | lbd | 91-95 | f | 6 | x | x | x | x |  |
| 24 | non-lbd | ad hs | 96-100 | f | 14 | x | x | x | x | x |
| 25 | non-lbd | ad hs | 86-90 | f | 8 | x | x | x | x | x |
| 26 | non-lbd | ad psp | 71-75 | f | 21 | x |  | x |  | x |
| 27 | non-lbd | ad | 61-65 | m | 5 | x | x | x | x | x |
| 28 | non-lbd | ad | 86-90 | m | 6 | x | x | x | x | x |
| 29 | non-lbd | ad | 91-95 | f | 8 | x | x | x | x | x |
| 30 | non-lbd | ad | 81-85 | f | 24 | x | x |  | x | x |
| 31 | non-lbd | ad | 76-80 | m | 9 | x | x | x | x | x |
| 32 | non-lbd | ad | 81-85 | f | 23 | x |  | x |  | x |
| 33 | non-lbd | hs | 86-90 | m | 6 | x |  | x | x |  |
| 34 | non-lbd | dooo | 91-95 | f | 16 | x | x |  | x | x |
| 35 | non-lbd | mii | 81-85 | m | 66 | x | x | x | x | x |
| 36 | non-lbd | nothing | 76-80 | m | 46 | x | x | x | x | x |
| 37 | non-lbd | control | 86-90 | m | 19 | x | x | x | x |  |
| 38 | non-lbd | nothing | 81-85 | m | 3 | x | x | x | x |  |
| 39 | non-lbd | control | 26-30 | m | 35 | x | x | x | x |  |
| 40 | non-lbd | nothing | 91-95 | f | 12 | x | x | x | x |  |
| 41 | non-lbd | nothing | 56-60 | m | 7 | x | x | x | x |  |
| 42 | non-lbd | nothing | 86-90 | m | 5 | x | x | x | x |  |

Abbreviations: inf = infarct, m = male, f = female, lbd = lewy body disease, pd = parkinsons disease, pdd = parkinsons disease dementia, ad = Alzheimer’s Disease, psp = Progressive supranuclear palsy, hs = hippocampal sclerosis, dooo = disease of the oldest old, mii = mild ischemic injury, Lrg = large, Fr = frontal, Midb/medul = midbrain/medulla, hippocamp = hippocampus,

### Real-Time Quaking-Induced Conversion (RT-QuIC) - Deparaffinization

A previously described protocol [16] was altered to deparaffinize tissue. For each sample, two 15μm sections were subjected to four 10-minute xylene washes with centrifugation (25°C for 5 minutes at 17,000 x g), fresh xylene overnight, and two more xylene washes in the morning. Two brief washes using 100% ethanol followed, with 10 seconds and 1 minute of vortexing respectively. Samples were shaken in a Thermomixer (Eppendorf ThermoMixer F1.5, Fisher Scientific 05-412-500) at 56°C for 3 minutes (300 rmp), centrifuged at 25°C for 5 minutes (17,000 x g), then subjected to three, 10-minute washes with 100% ethanol. During washes, samples were on a rocker and briefly vortexed every 2 minutes. Samples were spun between washes as described above. After the final wash, ∼80% of supernatant was removed, and the samples were left at 4°C overnight. The next day, samples were dried for 30min in a SpeedVac (Savant SPD1010, Thermo Fisher Scientific) set to 10 Torr/minute and ambient temperature.

### RT-QuIC - Tissue Processing

Samples were processed based on dry tissue weight as previously described [2, 16, 18] in TSB (pH 8.0, 500mM TRIS, 0.08% SDS, protease inhibitor (MilliporeSigma 11697498001, concentration per vendor). 500μl of TSB and fifteen, 0.7mm zirconia grinding beads (BioSpec 11079107ZX) were added to sample tubes. Samples were rocked for 30 seconds and vortexed for 30 seconds. Tubes were filled halfway with equal parts 0.7mm zirconia grinding beads and 0.4mm zirconia grinding beads (OPS Diagnostics BLBZ 400-250-37) and the remainder of the tube was filled with TSB. Samples underwent ten rounds of bead-beating (Mini-Beadbeater-Plus, BioSpec 1018), each round was 1 minute of homogenization and 1 minute in an ice water bath. Next, three 5-minute freeze/thaw cycles were performed by alternating samples between a -80°C ethanol bath and a 37°C water bath. Samples were then rocked for 10 minutes with intermittent vortexing, centrifuged at 25°C for 5 minutes (17,000 x g), old TSB was replaced, and then rocked for another 10 minutes. A 10% w/v homogenate in TSB was prepared in 0.2ml tubes. Shaking and heating of samples was performed at 90°C (1500rmp) for 20 minutes (Eppendorf Thermomixer F1.5, Fisher Scientific 05-412-500), then again at 60°C for another 90 minutes. Samples were briefly vortexed and then sonicated at 15°C using a cup-horn attachment and chiller (Qsonica Q700A-110, 431C2, 445, and 4905-110) (amp 30, on 3 seconds, off 2 seconds, 15 minutes of sonication). Samples were clarified 3-4 times by centrifugation at 4°C for 5 minutes (5000 x g) and stored at -80°C.

### RT-QuIC - Assay

Previously published RT-QuIC protocols [2, 16, 18] were adapted for FFPE tissue. Each well of a Nunc black, clear-bottomed, 96-well plate (Thermo Fisher Scientific 265301) was loaded with six 0.8 mm silica beads (OPS Diag BLBG 800-200-03). Reaction buffer consisted of 40 mM NaPO4, 170 mM NaCl, 20um ThT (filtered with a 0.22um syringe filter (MilliporeSigma SLGPM33RS)), and 0.1 mg/ml recombinant monomer. The recombinant α-synuclein monomer (StressMarq catalog code SAA-100, batch# NPD-R091.30, NPD-R091.31, and NPD-R091.36) for brain tissue and Proteos RP-003 or Tau dGAE C322A monomer (StressMarq cat# SAA-500, batch# NPD-R145.04) for gastrointestinal tissue, was thawed, filtered with Amicon 100kDa, 0.5ml filters (MilliporeSigma UFC510024), and added prior to plating. For the α-synuclein assay, initial testing included tissue concentrations from 10^-3^ to 10^-5^ of GI tissue homogenate and 10^-1^ and 10^-2^ dilutions of brain tissue homogenate. For the tau assay, a 10^-3^ dilution was used for both GI and brain tissue homogenates. After testing, 10^-3^ of GI tissue and 10^-1^ of brain tissue was used. 2μl of sample and 98μl of reaction buffer were plated in each well. The assay was run in a FLUOstar Omega plate reader (BMG Labtech) at 42°C (gastrointestinal tissue) or 37°C (brain tissue) and consisted of intermittent shaking (1 minute, 400rpm (gastrointestinal tissue) or 700rpm (brain tissue), double orbital) and rest (1 minute) as well as plate reading every 45 minutes (bottom read, 450nm excitation, 480nm emission). A baseline of 20,000 relative fluorescence units (rfu) was used to set optic gain. Onset of aggregation/lag time was defined as the time fluorescence exceeded 4x baseline and mean lag times were calculated across four technical replicates for each sample. Excluded replicates were those with near-instantaneous aggregation; a known artifact in RT-QuIC. For the main manuscript, all technical replicates were used for quantitative comparisons. An exception was a demonstration in the supplemental of use of duodenum RT-QuIC data for conventional binary classification of LBD vs non-LBD, which was performed using the 3-of-4 replicates rule, where a case is called positive if three of four replicates show fluorescence 4x above a baseline threshold by a certain time and the minority slower aggregating technical replicates are ignored.

### Microscopy and Image analysis

Paraffin sections (6μm) were deparaffinized and stained using eosin/luxol fast blue (ELFB) to identify histologic features or used in immunostaining preparations. Immunostaining was done after heat- and citrate-based antigen retrieval and used antibodies to α-synuclein (MJFR1; Abcam; Catalog No. ab138501), tau (PHF1, kind gift from lab of Peter Davies and Aβ (4G8; BioLegend; Catalog No. 800701). Stains were developed using horseradish peroxidase (Vector ELITE HRP kit). Slides were digitized at 40x resolution on an automated brightfield slide scanner (Aperio GT 450; Leica Biosystems). Sections of FFPE midbrain and medulla were prepared as described above and photobleached for 96 hours to remove autofluorescence. Immunofluorescence staining was performed using antibodies against protein gene product 9.5 (PGP 9.5) (Sigma-Aldrich; Catalog No. AB1761-I) and Chromogranin A (1:200) in duodenum tissues and tyrosine hydroxylase (TH) (F-11; Santa Cruz Biotechnology; Catalog No. sc-25269), 8-hydroxy-guanosine (OHG-6) (N45.1; OriGene Technologies; Catalog No. AM20226PU-S), and PGP 9.5 in midbrain and medulla tissues. AlexaFluor secondary antibodies were applied (ThermoFisher; Catalog No. A-21244 and A-21137) and (Jackson ImmunoResearch Laboratories Inc.; 115545205) and washed in 4,6-diamidino-2-phenylindole (DAPI; Invitrogen; Catalog No. 62248).

Mounting media (Invitrogen; S369175) was added to tissue before cover slipping (Fisher Scientific; Catalog No. 12541021). Full slide scans at 20x resolution were performed on an automated scanner (Axioscan 7; Zeiss). Quantification of Lewy Bodies and degenerative foci of pigment in macrophages were performed by manual counting in ELFB-stained slides within six high-power fields. Histologic evaluation of autolysis in gastrointestinal specimens was performed by a single blinded gastrointestinal pathologist (GS) for each tissue layer (10 = complete, 0 = none) and the estimated proportion of nuclear basophilia, lamina propria disruption/hyalinization, crypt base loss and submucosal gland dilation/architectural disruption. Histologic evaluation of morphologic features in brain sections was blinded and performed by the same single pathologist rater (GS) in randomly selected high-power fields after initial training and ongoing QA by a board-certified neuropathologist (RW). For image analysis of pigment and immunostaining, whole slide image files were imported into image analysis software, Visiopharm (Versions 2025.02.2x64, Visiopharm A/S, Hørsholm, Denmark). ROIs were manually identified for gray matter, white matter, and substantia nigra. Image segmentation of pigment, Lewy pathology, tau pathology and amyloid pathology was performed through manual annotation of all pixels in a blinded manner by a single rater (AQ) in 10+ representative high-power fields followed by training an automated detector in Visiopharm that could be deployed across all fields of all slides. Results were spot checked in randomly selected slides by a board-certified neuropathologist (RW). TH expression in the midbrain was measured by identifying neuron-associated pixels using whole-slide intensity thresholding of PGP 9.5 and OHG-8 (same threshold for all subjects) and mean neuronal TH intensity was normalized to PGP 9.5.

### Statistical Analysis

Statistical analysis was performed in RStudio (Posit Software, Boston, MA). Groupwise lag times were compared using a student’s t-test and ANOVA (functions *t.test* and *aov*). Pearson correlation was calculated using linear regression (function *lm)* and outliers were identified using standardized residuals (function *rstandard)* greater than 3. Results are reported as Pearson’s r correlation coefficient. Results were evaluated for the entire cohort and then subsequently in just LBD and non-LBD subgroups. A secondary analysis divided LBD into those with (LBD+AD) and without (LBD-no-AD) Alzheimer’s disease co-pathology based on neuropathologist evaluation at the time of autopsy.

## RESULTS

### Baseline characteristics and tissue quality

There were no significant differences of baseline characteristics (**Table 1**, **sFigure 1**) between LBD and non-LBD in terms of age (77.3 ±7.4 vs. 80.2 ±15.7 years), sex (69.6 vs. 57.9 % male), and post-mortem interval (17 ±9.5 vs. 17.4 ±16.3 hours). Seven LBD subjects (30%) had concomitant neuropathologic diagnosis of AD pathology. GI tissue autolysis was present, typically confined to the mucosa, and was not significantly different between groups (**sFigure 1**).

### Validation of seeded aggregation assay in brain tissue versus corresponding neuropathology

Seeding in brain and GI tissues was measured and lag times are shown in **Figure 1A**. To validate seeding against neuropathology, brain seeding was compared to neuropathologic features (**sFigure 2**). In the LBD group, seeding of α-synuclein in the frontal cortex was correlated with both α-synuclein immunohistochemistry (r = 0.63) and Lewy bodies in the substantia nigra (r = 0.51). Seeding of tau-dGAE was correlated in the non-LBD group with Braak tau stage in the frontal cortex (r = 0.73) and hippocampus (r = 0.79).

**Figure 1.**
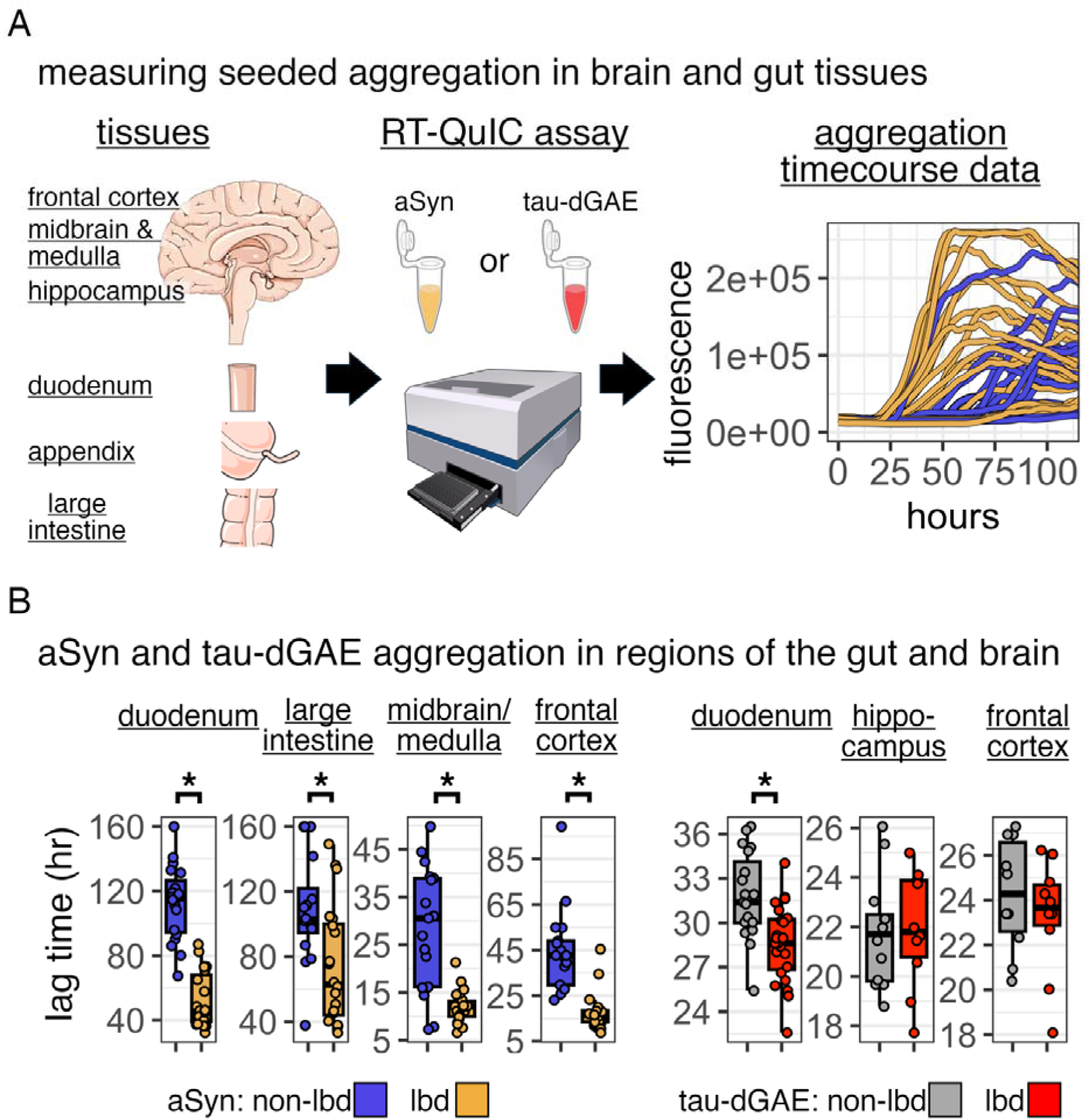
α-synuclein (aSyn) and tau-dGAE seeded aggregation is increased in the gastrointestinal tract and brain in LBD versus other cases. A: Project workflow performing RT-QuIC assays on gut and brain tissues and example timecourse fluorescence data. B: Aggregation lag times in LBD (gold and red) versus non-LBD cases (blue and gray) for α-synuclein and tau-dGAE. Mean of four technical replicates per patient. * p < 0.05

### Seeded aggregation of α-synuclein and tau-dGAE in the gastrointestinal tract

To test the dependency of gut seeding on the presence of neurodegenerative disease, seeding in gastrointestinal tissues was compared between LBD and non-LBD groups (**Figure 1B**). Seeding in duodenal tissues was significantly faster in LBD versus non-LBD for α-synuclein (51.8 ± 16.8 vs 112.6 ± 23.0 hours) and tau-dGAE (28.5 ± 2.6 vs 31.7 ± 2.9 hours) (**Figure 1B**) and there were progressively smaller albeit-still-significant groupwise differences for α-synuclein seeding in the appendix (63.7 ± 30.9 vs 105.9 ± 28.9 hours) and large intestine (74.3 ± 36.5 vs 108.7 ± 33.5) (**sFigure 3**). Duodenal α-synuclein seeding was correlated with duodenal tau-dGAE seeding in all subjects (r = 0.54) and just the LBD subgroup (r = 0.5) (data not shown). Seeded aggregation in gastrointestinal tissue was not correlated with participant age, post-mortem interval, atherosclerosis, or autolysis (**sFigure 4**).

As an aside, to demonstrate the use of seeding as a binary test in this specimen type, the fastest 3-of-4 replicates rule was used where a positive test has 3 of 4 technical replicates that aggregate before a determined time. Duodenal α-synuclein seeding showed sensitivity of 91.3% and specificity of 100% for binary classification of LBD versus non-LBD (**sFigure 5**). Additional testing was not performed including testing of earlier disease stage, performing endpoint dilution, and blinded external validation.

### Seeded aggregation in the brain versus seeded aggregation in the gastrointestinal tract

To compare seeding in the gut and brain, lag times for seeding in the brain were measured and then correlated to seeding in gastrointestinal tissues Seeded aggregation was measured in various brain regions and compared between LBD and non-LBD (**Figure 1B**). For α-synuclein, lag times were shorter in LBD for the frontal cortex (18.0 ± 8.6 vs 44.4 ± 18.4 hours) and midbrain/medulla (12.0 ± 3.3 vs 28.4 ± 13.0 hours). For tau-dGAE, lag times were not different between LBD and non-LBD in frontal cortex (23.2 ± 2.5 vs 24.2 ± 2.5 hours) or hippocampus (21.9 ± 2.3 vs 21.7 ± 2.2 hours).

Seeding in the brain was compared to seeding in gastrointestinal tissues through linear correlation (**Figure 2**). For α-synuclein, the duodenum was positively correlated with frontal cortex for all subjects (r = 0.75) and borderline for both LBD (r = 0.42, p = 0.079) and non-LBD (r = 0.43, p = 0.079) subgroups. Duodenum was correlated with midbrain/medulla for all subjects (r = 0.55) but only borderline for LBD (r = 0.35, p = 0.10). The large intestine was more strongly correlated with midbrain/medulla for all subjects (r = 0.63) and the non-LBD subgroup (r = 0.64). For the LBD subgroup there was range restriction of assay results (mean lag time = 12.0 ± 3.3 hours) and ten-fold dilution of midbrain/medulla demonstrated increased inter-subject variability (mean lag time = 48.8 ± 14.6 hours). At this lower input concentration of midbrain/medulla tissue, significant correlation with α-synuclein seeding in the large intestine was seen for the LBD subgroup (**Figure 2**, dark orange, r = 0.61) in addition to all subjects (r = 0.61) and the non-LBD subgroup (r = 0.40).

**Figure 2:**
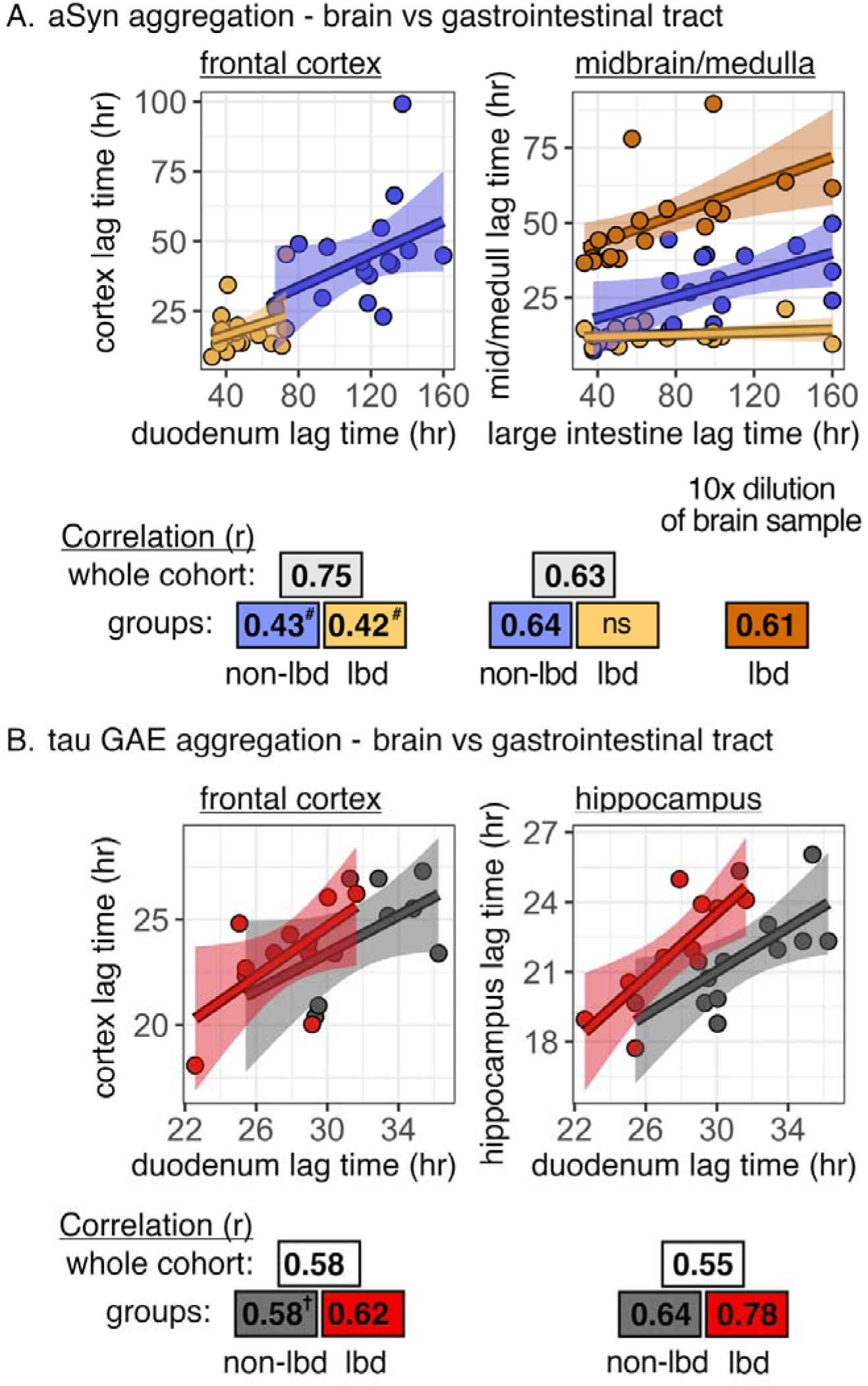
Correlation of α-synuclein (aSyn) and tau-dGAE aggregation in the brain with aggregation of the same proteins in the duodenum and large intestine. A: Correlation of α-synuclein lag times from the fro tal cortex (left) and midbrain/medulla (right) compared to the duodenum and large intestine in patients with (gold) and without (blue) LBD. B: Correlation of tau-dGAE aggregation lag times from brain regions with aggregation lag times from the duodenum. Linear regression (lines, confidence intervals), Pearson r values (p < 0.05), ns = not significant, # p = 0.079, † p = 0.073

For tau-dGAE seeding in brain versus gastrointestinal tissues, duodenum was correlated with hippocampus (r = 0.55) and frontal cortex (r = 0.58). In the LBD subgroup, duodenum was correlated with hippocampus (r = 0.78) and frontal cortex (r = 0.62). In non-LBD, duodenum correlated with hippocampus (r = 0.64) and borderline for frontal cortex (r = 0.58, p = 0.07).

Seeding across brain and gut and across α-synuclein and tau-dGAE were compared and there was one positive correlation for tau-dGAE in the duodenum and α-synuclein in the cortex (r = 0.44) and a negative correlation for just LBD between α-synuclein in the large intestine and tau-dGAE in the cortex (r = -0.63).

### Brain Lewy pathology versus α-synuclein seeded aggregation in the gastrointestinal tract

To measure how seeding of α-synuclein in the gut relates to central LBD neuropathology, histologic features were counted in the brain and correlated to gut lag times. Lewy pathology was measured using quantification of immunostained α-synuclein and manual identification of histologic Lewy bodies. The LBD group had markedly elevated α-synuclein-positive pixels in the substantia nigra (0.035 ± 0.023 vs 0.0022 ± 0.0019 % tissue area) and the dorsal motor nucleus of the vagus (0.0026 ± 0.0023 vs 0.0000011 ± 0.0000045 % tissue area) and the LBD group had Lewy bodies in the ventrolateral substantia nigra (0.34 ± 0.39 vs 0 ± 0 per high-power field) and medial substantia nigra (0.27 ± 0.34 vs 0 ± 0 per high-power field).

Lewy pathology in the LBD group was compared to gastrointestinal seeding (**Figure 3A**). Lewy bodies in the medial substantia nigra correlated with α-synuclein seeding in both the duodenum (r = -0.45) and borderline for large intestine (r = -0.38, p = 0.09). α-synuclein-positive pixels in the DMV (r = -0.50) were correlated with α-synuclein seeding just in the large intestine. No correlation was seen with Lewy bodies in the ventrolateral substantia nigra, and no correlation was seen between tau-dGAE seeding and Lewy pathology.

**Figure 3:**
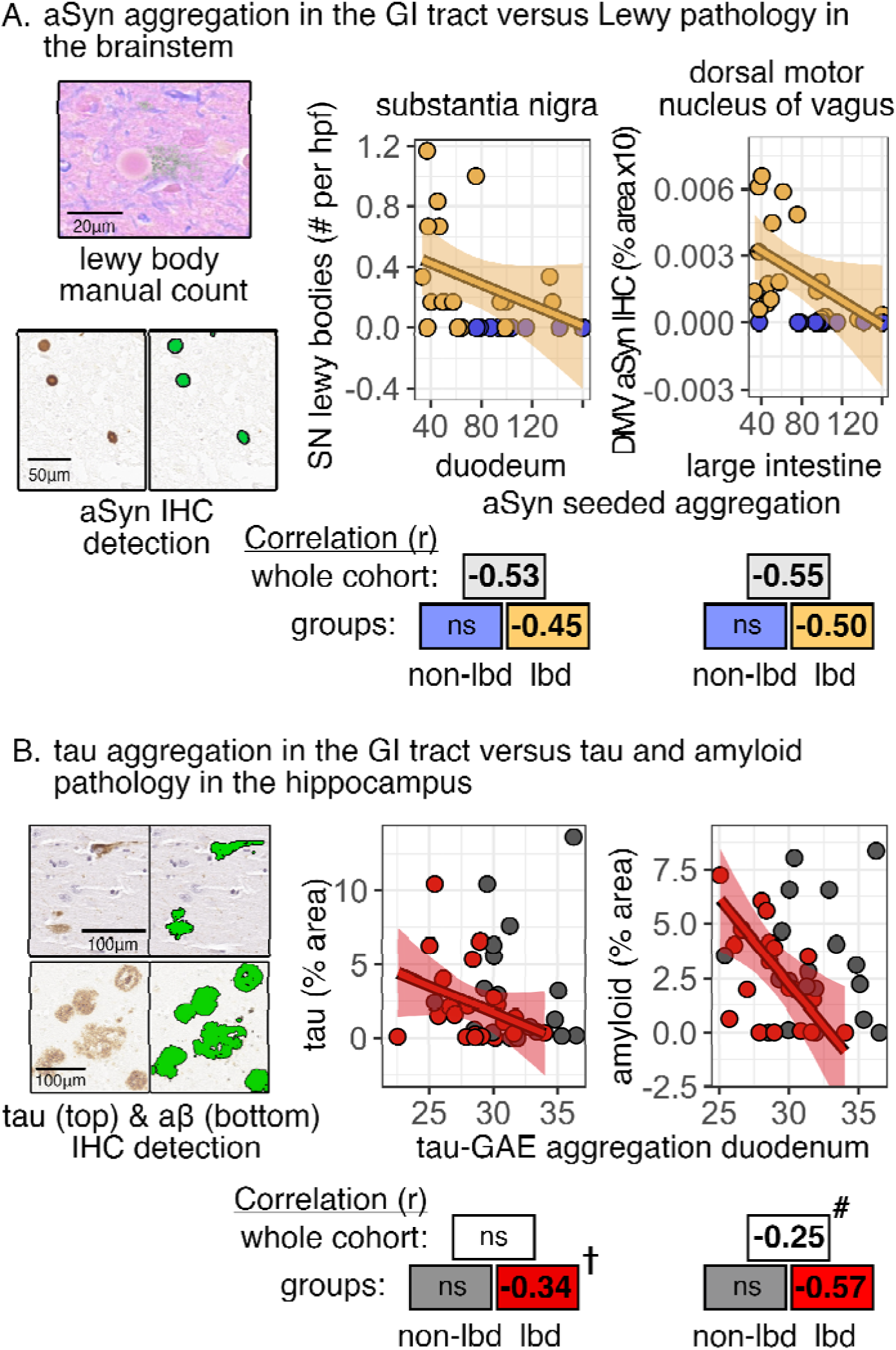
Comparison of lewy and amyloid pathology in the brain with α-synuclein (aSyn) and tau-dGAE aggregation in the gastrointestinal tract. A: Manual count of histologic Lewy bodies (eosinophilic inclusion, left image) and quantification of immunostained α-synuclein (green, detection via image analysis, left image) and correlation to α-synuclein aggregation lag times in duodenum and large intestine from participants with (gold) and without (blue) LBD. Panel B: Quantification of immunostained amyloid-β (aβ) and tau pathology (green, detection via image analysis, left image) in hippocampus and correlation to tau-dGAE aggregation lag times in duodenum. Linear regression (lines, confidence intervals), Pearson r values (p < 0.05), ns = not significant, † p = 0.12 and # p = 0.11.

### Tau and amyloid neuropathology compared to tau-dGAE seeded aggregation in the gastrointestinal tract and brain

To measure how seeding of tau in the gut relates to central tau and amyloid pathology, Lewy-related histologic features were counted in the brain and correlated to gut lag times. Tau and amyloid were measured in immunostained sections of hippocampus. There were no groupwise differences between LBD and non-LBD for amyloid-positive pixels (3.1 ± 3.2 vs 3.1 ± 2.6 % area, p = 0.9) or tau (2.3 ± 2.7 vs 3.1 ± 3.9 % area, p = 0.4).

Amyloid immunostaining in the hippocampus was correlated to tau-dGAE seeding in the duodenum for the LBD subgroup (r = 0.57) and borderline for the entire cohort (r = 0.25, p = 0.11) (**Figure 3B**). No correlation existed in the non-LBD group. Similar correlation, strongest in LBD, was present for tau-dGAE seeding in the hippocampus versus amyloid immunostaining in the hippocampus (**sFigure 7**). Tau immunostaining in the hippocampus was borderline correlated with tau-dGAE seeding in the duodenum just for the LBD subgroup (r = -0.34, p = 0.12).

No significant correlation was present between tau and amyloid in the hippocampus versus α-synuclein seeding in the duodenum or large intestine.

### Nigral neurodegeneration versus seeded aggregation in the gastrointestinal tract

To measure how seeding in the gut relates to central neurodegenerative pathology, nigral pathologic features were counted in the brain and correlated to gut lag times. Degeneration of dopaminergic neurons was measured in histologic sections of the substantia nigra by image analysis of total neuromelanin pigment and manual count of pigmented neurons and neuromelanin-laden macrophages. The LBD group had less total pigment in the substantia nigra (0.25 ± 0.18 vs 0.38 ± 0.19 % total SN area), fewer pigmented foci (586 ± 337.2 vs 1041 ± 400.6 no. foci), and fewer pigmented neurons (6.0 ± 3.0 vs. 9.1 ± 3.2 per high-power field). There were borderline increased neuromelanin-laded macrophages in the ventrolateral substantia nigra (3.7 ± 2.1 vs 2.9 ± 1.3 per high-power field, p = 0.19) and increased numbers but no groupwise differences in medial substantia nigra (4.0 ± 2.5 vs. 4.2 ± 2.7 per high-power field, p = 0.8). In the dorsal motor nucleus of the vagus, the LBD and non-LBD groups did not have significantly different numbers of pigmented neurons (1.5 ± 1.9 vs. 1.0 ± 1.5, p = 0.3) or neuromelanin-laded macrophages (2.1 ± 7.3 vs 1.3 ± 3.1, p = 0.6).

Seeding in the gastrointestinal tract was compared to histologic measures of neurodegeneration (**Figure 4A**). Nigral pigment loss correlated strongly with α-synuclein seeding in the large intestine (r = 0.76) and less so with the duodenum (r = 0.45, p < 0.05). For the large intestine this correlation was significant in subgroup analysis of both LBD (r = 0.67) and non-LBD (r = 0.65). Using partial correlation, findings were replicated after adjusting for subject age (**sFigure 6**). Findings were similar when using manual counts of pigmented neurons with strongest correlation to α-synuclein seeding in the large intestine (r = 0.56) that was significant in the LBD subgroup (r = 0.42) but not non-LBD (data not shown). Neuromelanin-laded macrophages in ventrolateral and not medial substantia nigra were correlated with the duodenum in the entire cohort (r = -0.37) and the non-LBD subgroup (r = -0.58). In the LBD subgroup, increased seeding speed in the large intestine was also correlated with decreased neuronal Tyrosine hydroxlase expression (r = 0.49) in immunolabeled neurons within the midbrain with the substantia nigra (**Figure 4A**).

**Figure 4:**
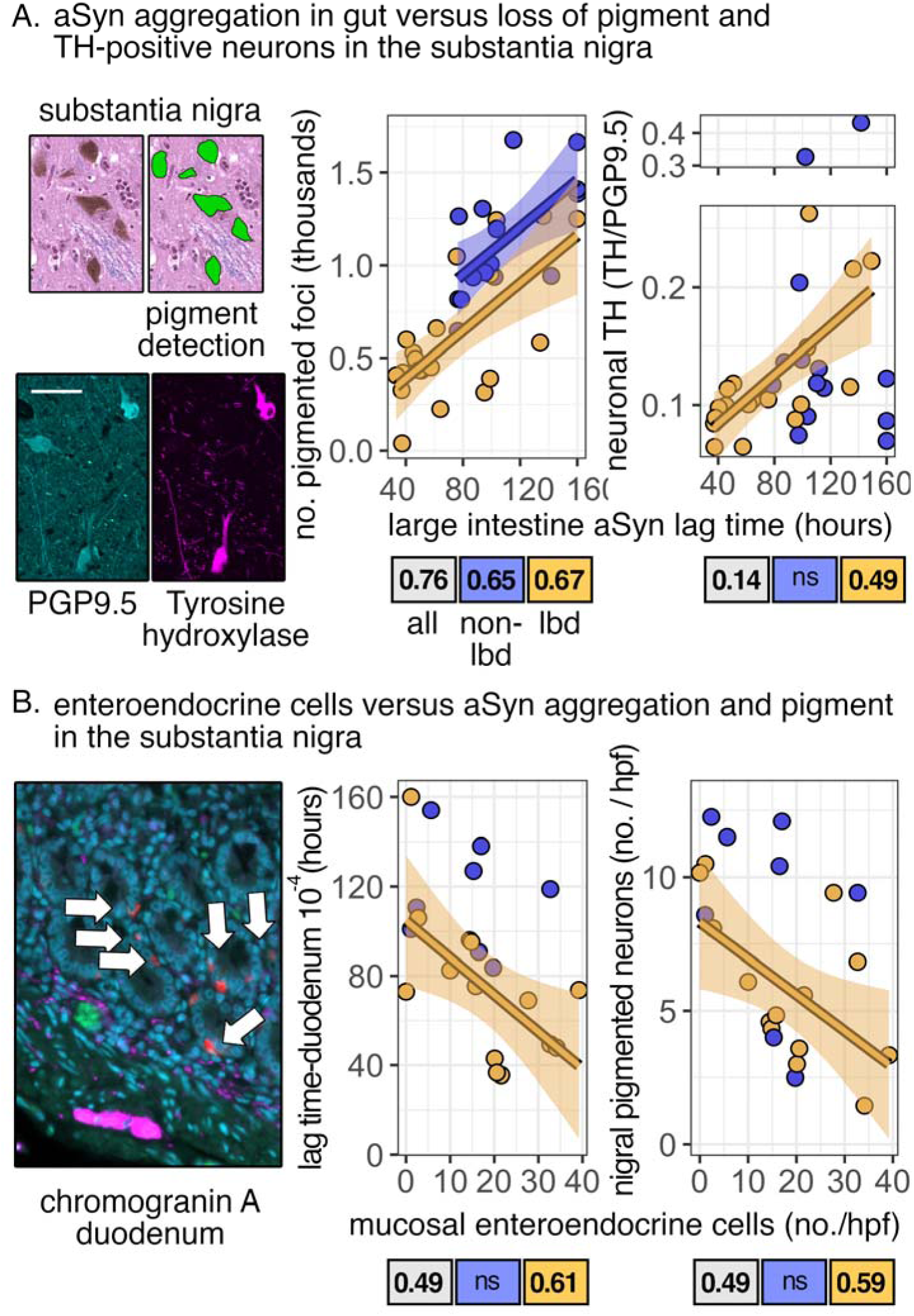
Comparison of neurodegeneration, α-synuclein (aSyn) seeding in the gut, and enteroendocrine cells in the duodenum. A: Left: Quantification of pigment in substantia nigra (middle graph) and tyrosine hydroxylase expression in PGP+OHG-8+ neurons (right graph) and correlation to α-synuclein lag times in large intestine (example images on left, inset = 100µm) in the entire cohort (gray) and the LBD (yellow) and non-LBD subgroups (blue). B: Manual count of duodenal enteroendocrine cells (white arrows) in a subset of cases using chromogranin-A and correlation to α-synuclein seeding in 10^-4^ duodenal tissue (middle graph) and nigral pigment (right graph). Example image shows duodenal mucosa (red = chromogranin-A, blue = DAPI, pink = PGP 9.5). Linear regression (lines, confidence intervals), Pearson r values (p < 0.05), ns = not significant.

### Mucosal enteroendocrine cells, seeded aggregation, and nigral neurodegeneration

To pilot test the relationship of seeding and pathology to an implicated cell type in the gut, enteroendocrine cells were counted in a subset of GI specimens and compared to seeding and neuropathology. In a subset of cases, enteroendocrine cells were immunolabeled and manually counted in the duodenal mucosa. There were no significant differences in counts of enteroendocrine cells in LBD versus non-LBD (18.2 ± 12.3 vs. 13.7 ± 10.4). When comparing counts of enteroendocrine cells to seeded aggregation of α-synuclein in the same duodenal tissue, results show increased enteroendocrine cells with faster seeded aggregation (r = 0.49 for entire cohort, r = 0.61 for LBD only) (**Figure 4B**). Note that a lower 10^-4^ concentration of duodenum tissue was used with more intersubject variability **(sFigure 3)**. Increased counts of enteroendocrine cells were also correlated with loss of pigment in the substantia nigra (r = 0.49 for entire cohort, r = 0.59 for LBD only) (**Figure 4B)**.

### Comparison of seeded aggregation in Lewy Body Disease with versus without Alzheimer’s co-pathology

To test the relationship of seeding to Alzheimer’s disease co-pathology, neuropathologist diagnosis was compared to lag times in the gut for both α-synuclein and tau. The LBD cohort was split based on the presence of concurrent Alzheimer’s disease pathology determined by neuropathologist diagnosis (LBD+AD) versus without (LBD-no-AD). No differences in seeding were present between LBD+AD versus LBD-no-AD in the gut or brain (**sFigure 8**). LBD-no-AD had borderline faster α-synuclein aggregation in the large intestine versus non-LBD (66.1 ± 31.4 vs 108.7 ± 33.5 hours, p = 0.08) and LBD+AD did not (83.9 ± 45.6 hours). In contrast, only LBD+AD had faster tau-dGAE aggregation in the duodenum versus non-LBD (27.8 ± 1.9 vs 31.7 ± 2.9 hours) and LBD-no-AD did not (28.9 ± 2.8 hours).

## DISCUSSION

The current study demonstrates seeded aggregation in the gut and brain along with histologic measures of neuropathology. Data show that seeding in the gastrointestinal tract mirrors central seeding, Lewy-, and AD co-pathology and mirrors neurodegeneration independent of aging and the presence of a synucleinopathy. Preliminary results show mucosal enteroendocrine cells provide overlapping data to seeded aggregation. Overall, the GI tract provides an accessible window for studies of pathogenesis and clinical biomarkers. Data using archival tissues opens the benefits of large well-characterized retrospective cohorts with neuropathologic assessment.

### Seeded aggregation of α-synuclein in the absence of Lewy Body Disease

Data here demonstrate that seeding in the periphery (gut) mirrors seeding in the brain regardless of the presence of LBD. The speed of seeding was faster in LBD but even in non-LBD controls (a mixture of healthy controls and non-LBD neurodegenerative disease) seeding in the gut still strongly correlated with the brain. There is limited prior data for comparison and technical differences. Two previous studies from experienced labs tested seeded aggregation in the gut of PD versus controls and found excellent specificity [13, 24]. Perhaps most importantly, the reaction time limit in these studies was shorter at 40 hours which data above suggest is too short to capture most non-LBD aggregation. In support of this, data from both previous studies showed occasional technical replicates and aggregate data where controls were beginning to aggregate at the end of their time window. Other technical variables may also play a role such as use of fresh/frozen tissue and different monomer protein substrate. In terms of the magnitude of association of peripheral and central seeding, when data did not have a restricted range there did not appear to be differences in the slope of linear regression between non-LBD and LBD. Testing of explicit subgroups such as Alzheimer’s disease and input of additional tissue concentrations is needed to further explore brain-gut seeding relationships.

### Seeded aggregation of α-synuclein in Lewy body disease

The finding in the current study demonstrating increased seeding of α-synuclein in LBD in the gut and association with severity of synucleinopathy aligns with prior studies using other measures and supports the use of seeding assays alongside the benefits of retrospective studies. Numerous (immuno-) histochemical studies have identified Lewy bodies and abnormal (phospho-)α-synuclein in gut nerve plexi and nerves [4, 9, 23, 25]. In these studies, gut staining was highly specific and weakly-to-moderately sensitive for LBD, with sensitivity improving in more developed and severe disease. Our lab’s efforts using α-synuclein immunofluorescence were unsuccessful to obtain specificity for LBD in GI nerves or enteroendocrine cells which others have ascribed to use of formalin fixative rather than paraformaldehyde-based fixatives (e.g. Zamboni’s) along with limited sampling [12]. For seeded aggregation there have been fewer studies but they show similar trends to histology where there is moderate sensitivity and high specificity for discerning LBD and the assay sensitivity increases with more advanced disease and larger/deeper tissue sampling [13, 24]. Our data also aligns with mapping of α-synuclein in the gut. The decreasing gradient of sensitivity in the current study from duodenum to appendix to large intestine aligns with the rostro-caudal gradient of synuclein seen by immunohistochemistry and positivity rates in other seeded aggregation assays [3, 15]. The concordance of data is encouraging for the use of seeded aggregation in archival tissues for retrospective studies ideally with paired neuropathologic assessment. Such studies can circumvent time course bottlenecks by spanning the decades of preclinical/prodromal disease for research on pathophysiology and neuroprotective treatments and spanning the decades of LBD progression for research on disease modification.

### Seeded aggregation of tau-dGAE in the gut and brain

Seeded aggregation of tau in the gastrointestinal tract has not been measured in human neurodegenerative disease but can be informed by limited corresponding data from animal studies and human data in the skin. Animal studies have shown pathologic tau in the enteric nervous system is related to models of neurodegeneration. In mouse and rat models of AD and aging, enteric neurons and myenteric plexi have been shown to harbor increased phosphorylated tau, total tau, and Aβ with a subset colocalized to α-synuclein [14, 19]. Furthermore, tau was suggested in mouse models to be induced by gut inflammation and travel rostrally along the vagus in association with worse learning and memory that is rescued by vagotomy [8]. In humans, tau is expressed in enteric neurons but immunohistochemical studies of its relationship with neurodegenerative disease have been negative or inconclusive [11]. Prior human studies of seeded aggregation in the periphery have used skin and generally focused on discerning controls from pure and mixed tauopathies such as progressive supranuclear palsy (PSP), corticobasal degeneration (CBD), and Alzheimer’s Disease with moderate-to-excellent accuracy [10, 26]. A recent detailed study included cases of LBD and found elevated tau seeding in the skin, agreeing with data here showing elevated tau seeding in the gastrointestinal tract in LBD. An interesting difference in findings is that tau seeding in the skin did not show a relationship to AD co-pathology in the brain. This study included neuropathologic evaluation and reported on a subset of four DLB participants with AD co-pathology, but seeding was not visibly elevated in four participants within the DLB subgroup and was also not elevated in DLB versus PD. This gut versus skin difference in the relationship of tau seeding to central AD co-pathology could have entirely technical or biologic explanations such as different tau monomers and LBD enrollment or differences in the enteric and cutaneous nervous systems [1, 17, 20–22]. For example, the tau-dGAE monomer may be an ideal substrate to model Alzheimer’s co-pathology given 1) its isolation from the core of paired helical filaments in neurofibrillary tangles, 2) aggregation without the need for anionic additives like heparin, and 3) prion-like seeding of full-length tau into disease-associated conformations [17]. Further human studies are needed to directly compare seeding at different peripheral sites.

### Enteroendocrine cells are a possible link to seeding

Current data and previous studies suggest enteroendocrine cells are a potentially useful indicator of enteric neuropathology. Enteroendocrine cells are a rare lining cell of the gastrointestinal tract and play a combined role sampling luminal contents, making and secreting hormones, and interacting with the enteric nervous system. Enteroendocrine cells, like seeding, are potentially poised for clinical translation as these are mucosal-based cells that can be routinely biopsied. Transgenic- and exposure-based mouse models suggest that these cells have similar properties to neurons and synapse basally with enteric neurons where in PD models cross-sectional data suggest they transynaptically transfer α-synuclein to the enteric nervous system [6]. Human data shows expression of (phospho-)α-synuclein and (phospho-)tau in enteroendocrine cells [5, 7] and diseases such as inflammatory bowel disease are associated with PD risk and have increased enteroendocrine cells [27]. As noted above, we attempted α-synuclein immunohistochemistry in GI tissue and did not see LBD specificity, possibly due to use of formalin and not paraformaldehyde-based fixative [12]. For cell counts of enteroendocrine cells, early data here are in line with these prior findings by showing a quantitative link between counts of enteroendocrine cells and seeded aggregation.

### Limitations

An important limitation of the current study is a lack of additional clinical measurements such as neurologic and gastrointestinal symptoms, disease duration, and standardized rating instruments (e.g. UPDRS, MOCA). The current study contained diagnosis, baseline patient characteristics, and neuropathologic evaluation and for the LBD group, pathologic staging is Braak stage 3+ which generally corresponds with clinical manifestation (tremor, slowness, stiffness, and/or postural instability). Explicit measurement of these and other ancillary data (e.g. UPSIT smell test, cardiac scintigraphy, DatScan, LRRK2/GCA genotype) will help connect peripheral seeding to studies of pathogenesis and in vivo data for clinical research. Encouragingly, findings are independent of aging, which typically is one of the largest effect confounders in neurodegenerative disease research. Another limitation was the lack of testing smaller superficial biopsy specimens. Full thickness specimens were used in the current study to sample more of the enteric nervous system and maximize sensitivity for quantitative comparisons. Use of GI seeding in vivo and longitudinally will require studies of a feasible smaller biopsy specimen that is limited to mucosa and submucosa. Data here show enteroendocrine cells located in the mucosa may offer a superficial surrogate for biopsy. For studies outside of screening biopsies, full thickness and deeper GI tissues are often available as they are some of the most common larger and deeper sampling procedures performed in the healthcare system (e.g. sleeve gastrectomy, appendectomy, colorectal resections, rectal suction biopsy).

Overall, the current study demonstrates an underlying quantitative mirroring of seeded aggregation in the gastrointestinal tract and central nervous system for both α-synuclein and tau (modeled in **Figure 5**). This relationship is present regardless of diagnosis and correlated with central neuropathology suggesting it is not sufficient for disease but can serve as a quantitative biomarker of LBD and AD co-pathology. Early data also show the possible utility of mucosal enteroendocrine cells to provide similar information about central neuropathology. Data above collectively show the potential of gastrointestinal tissues to inform clinical biomarkers and basic disease pathogenesis.

**Figure 5:**
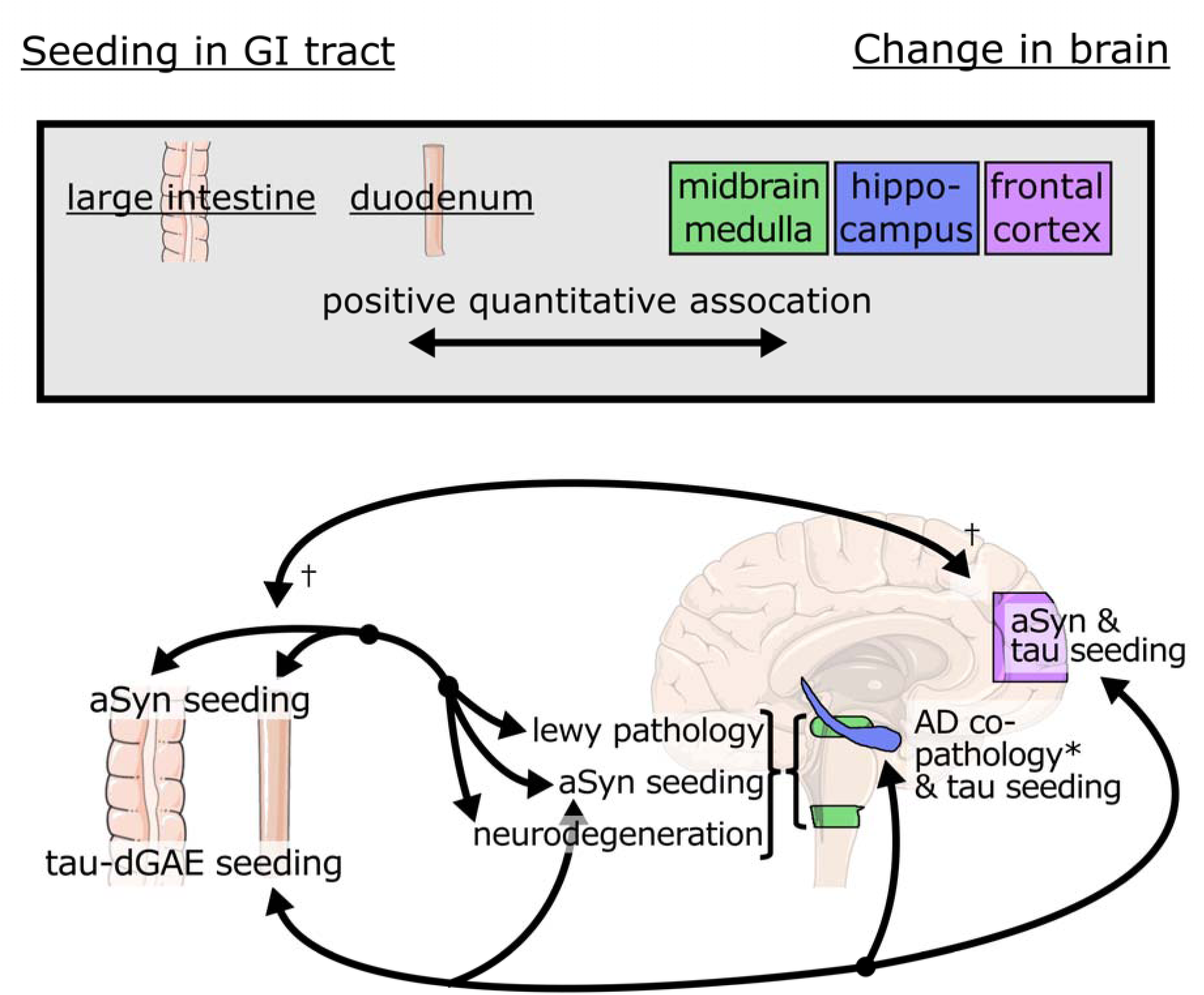
Gastrointestinal seeding as a window into neurodegeneration. Diagram shows significant positive (black arrows) correlation between seeding of α-synuclein and tau-dGAE in the duodenum and large intestine with seeding and pathologic features in the brain. * Association identified in LBD group and not identified in non-LBD - possibly from low numbers of neurodegenerative non-synucleinopathies (e.g. AD, PSP). † Association was borderline significant, possibly from restricted range of assay at tested tissue concentrations.

## Data Availability

All data produced in the present study are available upon reasonable request to the authors

## SUPPLEMENT

**sTable 1:**

| No. | Group | Athero-sclerosis | Neuritic Plaques | Diffuse Plaques | Braak Stage | Lewy Bodies Midbrain | Lewy Bodies Limbic | Lewy Bodies Neocortex |
|---|---|---|---|---|---|---|---|---|
| 1 | LBD | 1 | 2 | 3 | 6 | 1 | 1 | 1 |
| 2 | LBD | 1 | 2 | 3 | 6 | 1 | 1 | 1 |
| 3 | LBD | 1 | 2 | 3 | 6 | 1 | 1 | 1 |
| 4 | LBD | 0 | 2 | 2 | 5 | 1 | 1 | 1 |
| 5 | LBD | 1 | 1 | 2 | 6 | 1 | 1 | 1 |
| 6 | LBD | 2 | 2 | 2 | 6 | 1 | 1 | 1 |
| 7 | LBD | 0 | 2 | 2 | 6 | 1 | 1 | 1 |
| 8 | LBD | 2 | 2 | 3 | 6 | 0 | 1 | 1 |
| 9 | LBD | 2 | 0 | 0 | 3 | 1 | 1 | 1 |
| 10 | LBD | 0 | 0 | 1 | 0 | 1 | 1 | 1 |
| 11 | LBD | 0 | 2 | 2 | 4 | 1 | 1 | 1 |
| 12 | LBD | 0 | 1 | 1 | 4 | 1 | 1 | 1 |
| 13 | LBD | 0 | 0 | 0 | 3 | 1 | 1 | 1 |
| 14 | LBD | 0 | 0 | 2 | 1 | 1 | 1 | 1 |
| 15 | LBD | 0 | 0 | 2 | 1 | 1 | 1 | 1 |
| 16 | LBD | 0 | 0 | 3 | 0 | 1 | 1 | 1 |
| 17 | LBD | 1 | 0 | 0 | 1 | 1 | 1 | 1 |
| 18 | LBD | 2 | 3 | 2 | 0 | 1 | 1 | 1 |
| 19 | LBD | 0 | 0 | 0 | 1 | 1 | 1 | 1 |
| 20 | LBD | 1 | 0 | 0 | 3 | 1 | 1 | 1 |
| 21 | LBD | 2 | 0 | 0 | 4 | 1 | 1 | 0 |
| 22 | LBD | 0 | 0 | 0 | 2 | 1 | 0 | 0 |
| 23 | LBD | 2 | 2 | 2 | 5 | 1 | 0 | 0 |
| 24 | Non-LBD | 2 | 1 | 2 | 4 | 0 | 0 | 0 |
| 25 | Non-LBD | 0 | 2 | 2 | 5 | 0 | 0 | 0 |
| 26 | Non-LBD | 0 | 1 | 2 | 4 | 0 | 0 | 0 |
| 27 | Non-LBD | 1 | 3 | 3 | 6 | 0 | 0 | 0 |
| 28 | Non-LBD | 2 | 1 | 2 | 3 | 0 | 0 | 0 |
| 29 | Non-LBD | 2 | 1 | 2 | 4 | 0 | 0 | 0 |
| 30 | Non-LBD | 1 | 3 | 2 | 6 | 0 | 0 | 0 |
| 31 | Non-LBD | 2 | 2 | 2 | 6 | 0 | 0 | 0 |
| 32 | Non-LBD | 0 | 2 | 3 | 6 | 0 | 0 | 0 |
| 33 | Non-LBD | 1 | 0 | 0 | 2 | 0 | 0 | 0 |
| 34 | Non-LBD | 0 | 1 | 3 | 4 | 0 | 0 | 0 |
| 35 | Non-LBD | 0 | 1 | 1 | 2 | 0 | 0 | 0 |
| 36 | Non-LBD | 2 | 2 | 3 | 5 | 0 | 0 | 0 |
| 37 | Non-LBD | 0 | 0 | 1 | 2 | 0 | 0 | 0 |
| 38 | Non-LBD | 1 | 0 | 1 | 4 | 0 | 0 | 0 |
| 39 | Non-LBD | 0 | 0 | 0 | 0 | 0 | 0 | 0 |
| 40 | Non-LBD | 2 | 2 | 3 | 6 | 0 | 0 | 0 |
| 41 | Non-LBD | 0 | 3 | 2 | 6 | 0 | 0 | 0 |
| 42 | Non-LBD | 1 | 1 | 1 | 3 | 0 | 0 | 0 |

**sFigure 1:**
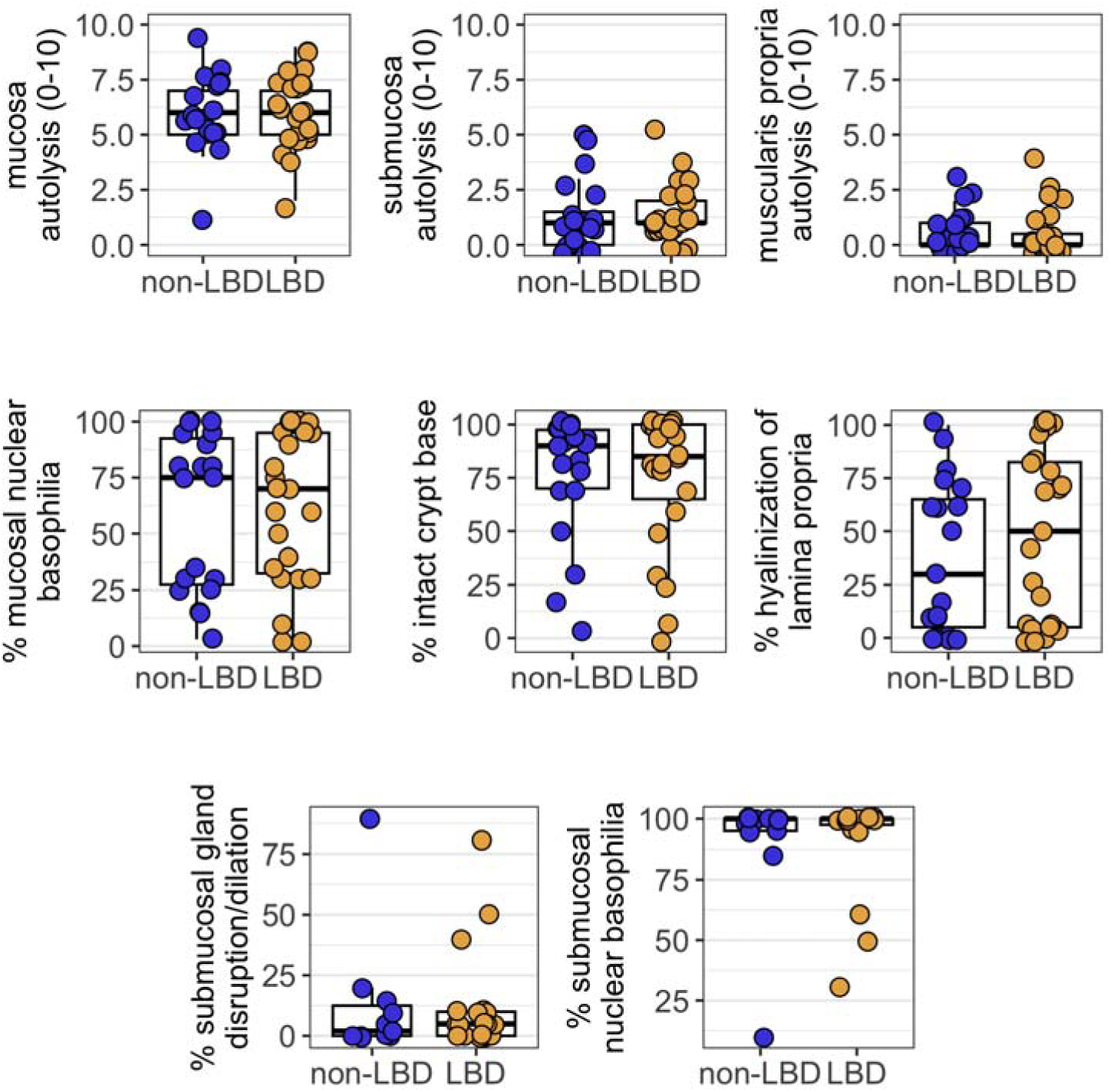
Autolysis in gastrointestinal tissue. Summary measures of autolysis (top row) and individual features in the mucosa (middle row) and submucosa (bottom row) measured by gastrointestinal pathology (GS) are shown for LBD (yellow) and non-LBD (blue) groups. No significant groupwise differences are present.

**sFigure 2:**
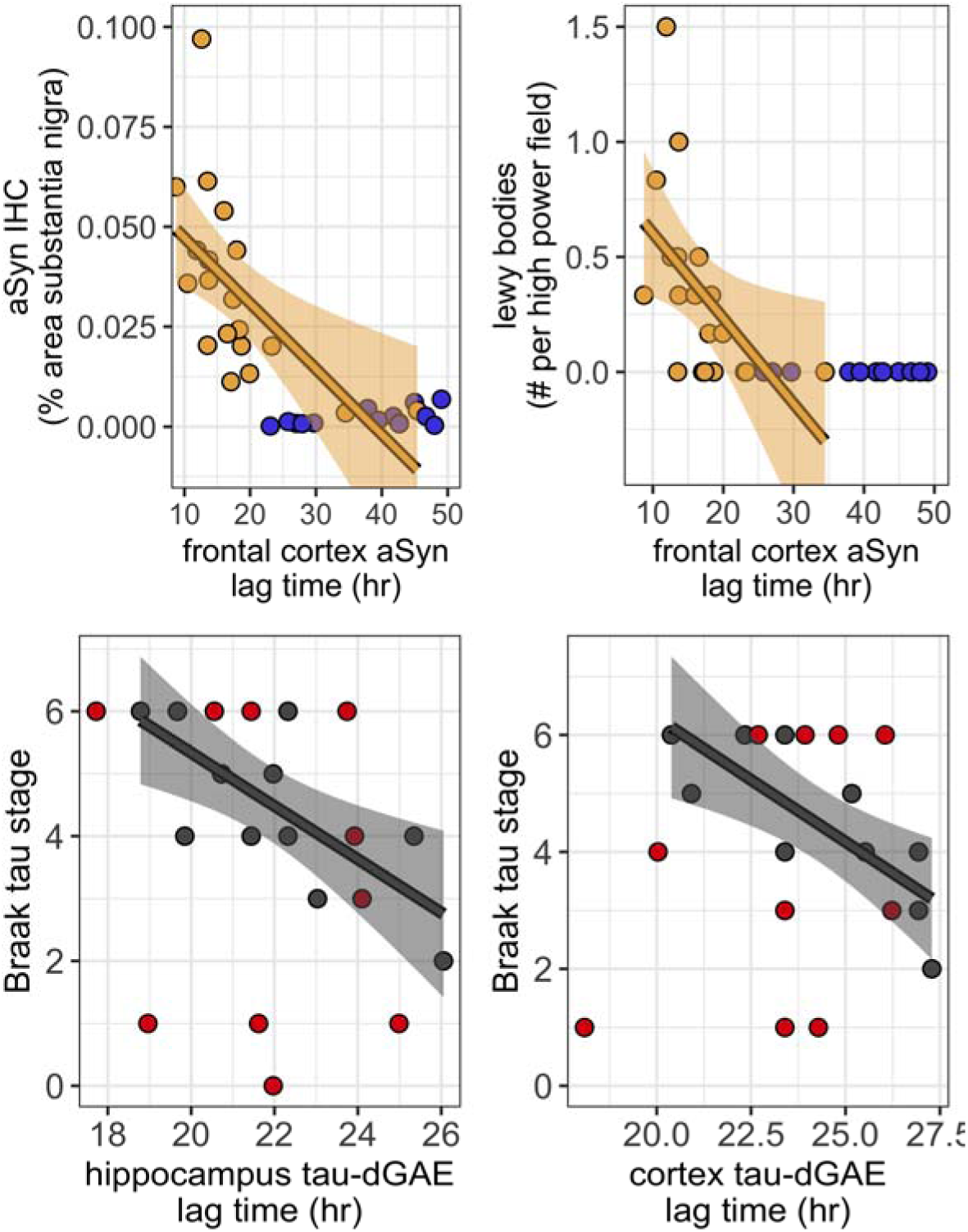
Correlation of α-synuclein and tau-dGAE aggregation in the brain with neuropathologic measures of Lewy and tau pathology. Top Panel: Correlation of α-synuclein lag times from the frontal cortex with quantity of α-synuclein + pixels (left) and manually counted Lewy bodies in random high-power fields (right) of the substantia nigra. Bottom Panel: Correlation a tau-dGAE aggregation lag times in hippocampus (left) and frontal cortex (right) with Braak tau stage. Lines indicate linear regression (lines, confidence intervals), p < 0.05. Note: panel A clips maximum lag time and does not show multiple non-LBD datapoints with long lag times in order to magnify the LBD group. These missing non-LBD datapoints lacked Lewy pathology.

**sFigure 3:**
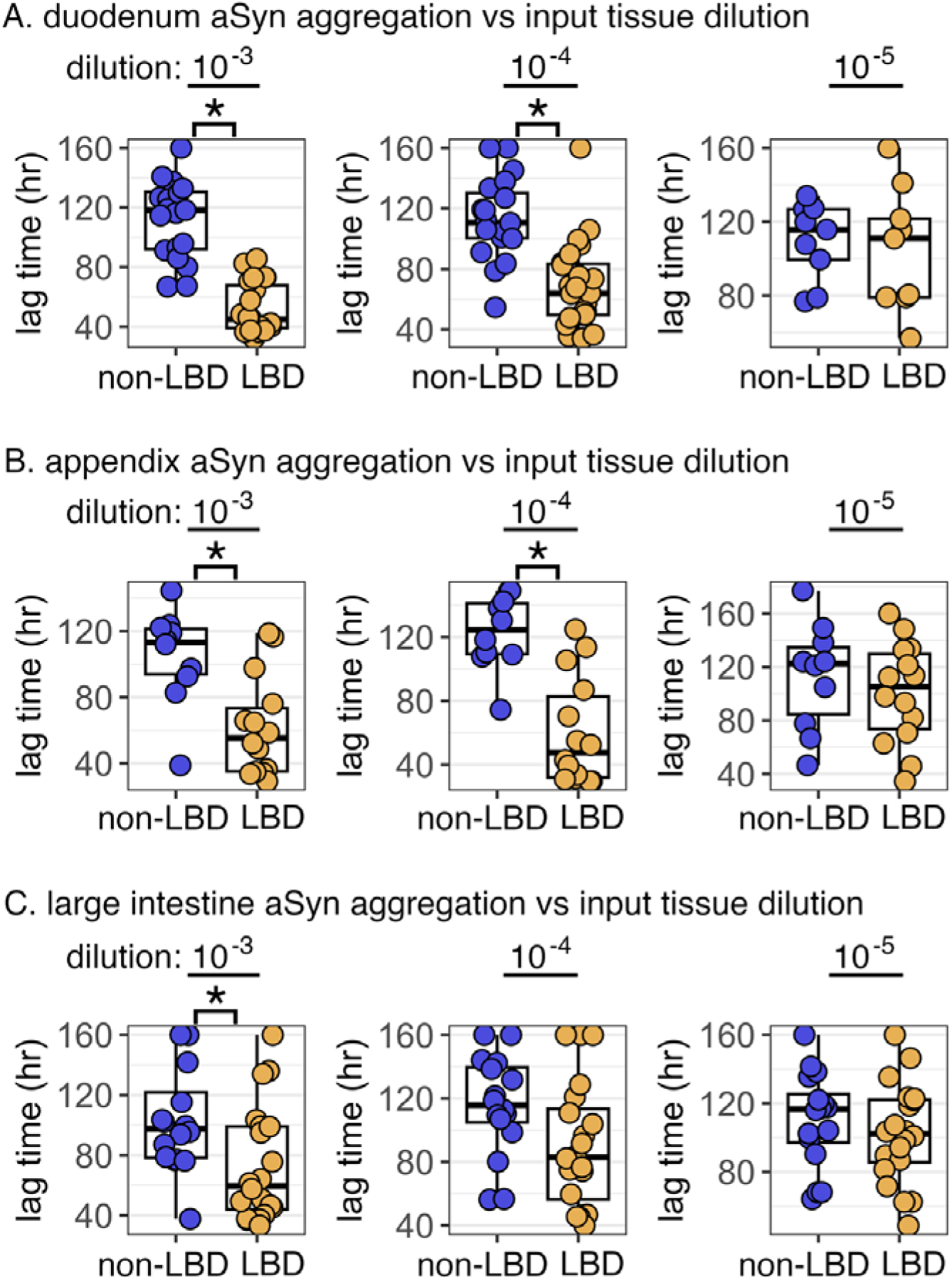
Tissue concentration versus seeded aggregation of α-synuclein in gastrointestinal tissues. Boxplot comparison of aggregation lag times between groups for α-synuclein in the duodenum (top), appendix (middle), and large intestine (bottom). * p < 0.05

**sFigure 4:**
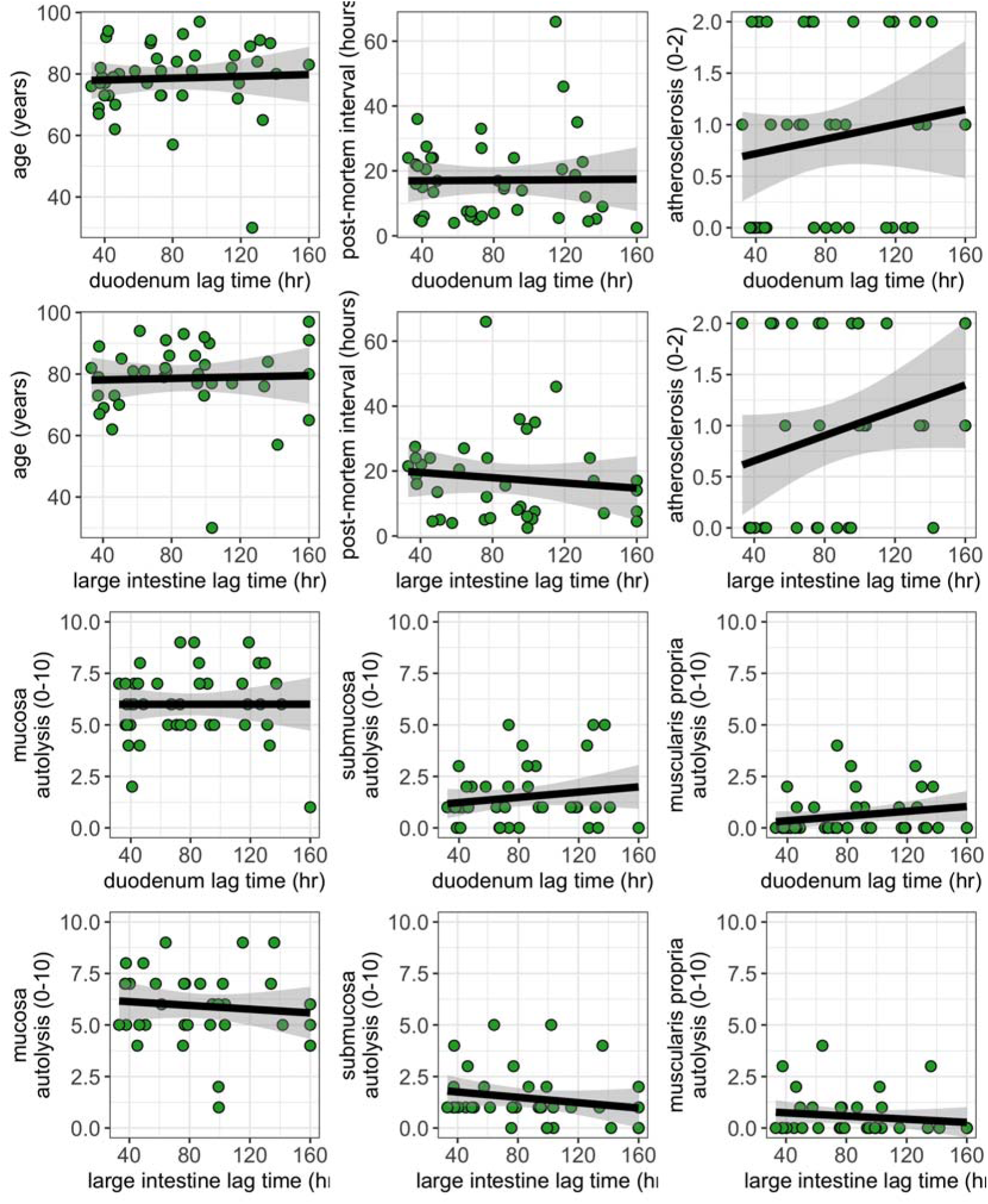
Comparison of baseline characteristics and tissue autolysis with α-synuclein seeded aggregation in the gut. Data are shown comparing baseline characteristics (age, post-mortem interval, atherosclerosis) and tissue autolysis in the mucosa, submucosa, and muscularis propria versus seeded aggregation of aSyn in the duodenum and large intestine. No significant or borderline correlations were present.

**sFigure 5:**
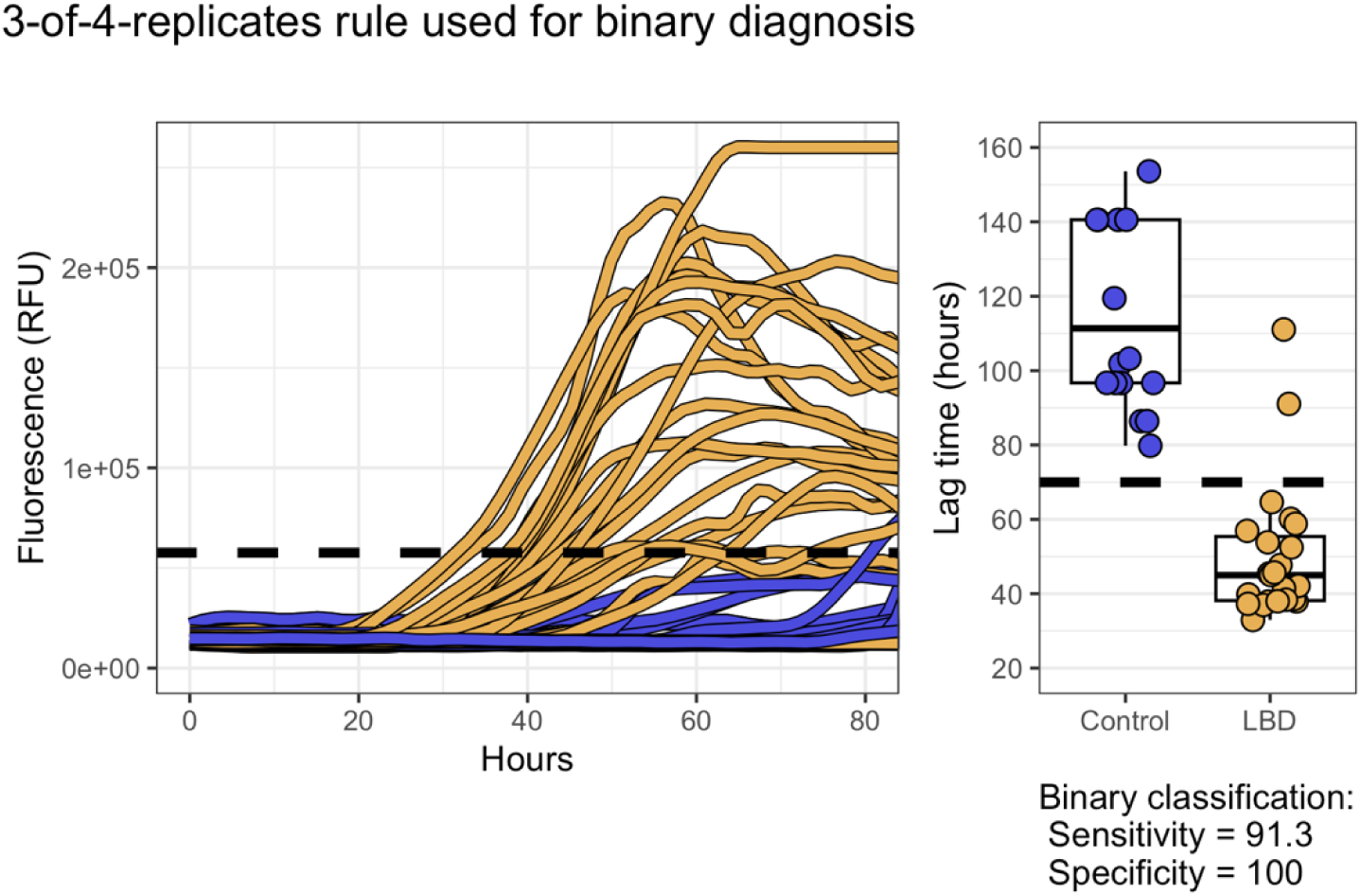
Binary classification for LBD with α-synuclein aggregation in the duodenum. Data show third fastest technical replicate, reflecting three-of four-technical-replicates rule. Positive binary classification defined as aggregation within ∼75s duration. Diagnostic performance is Sensitivity 91.3% and Specificity 100%. Endpoint dilution, earlier synucleinopathy cases, and external validation is not performed and is required to determine true binary diagnostic test performance.

**sFigure 6:**
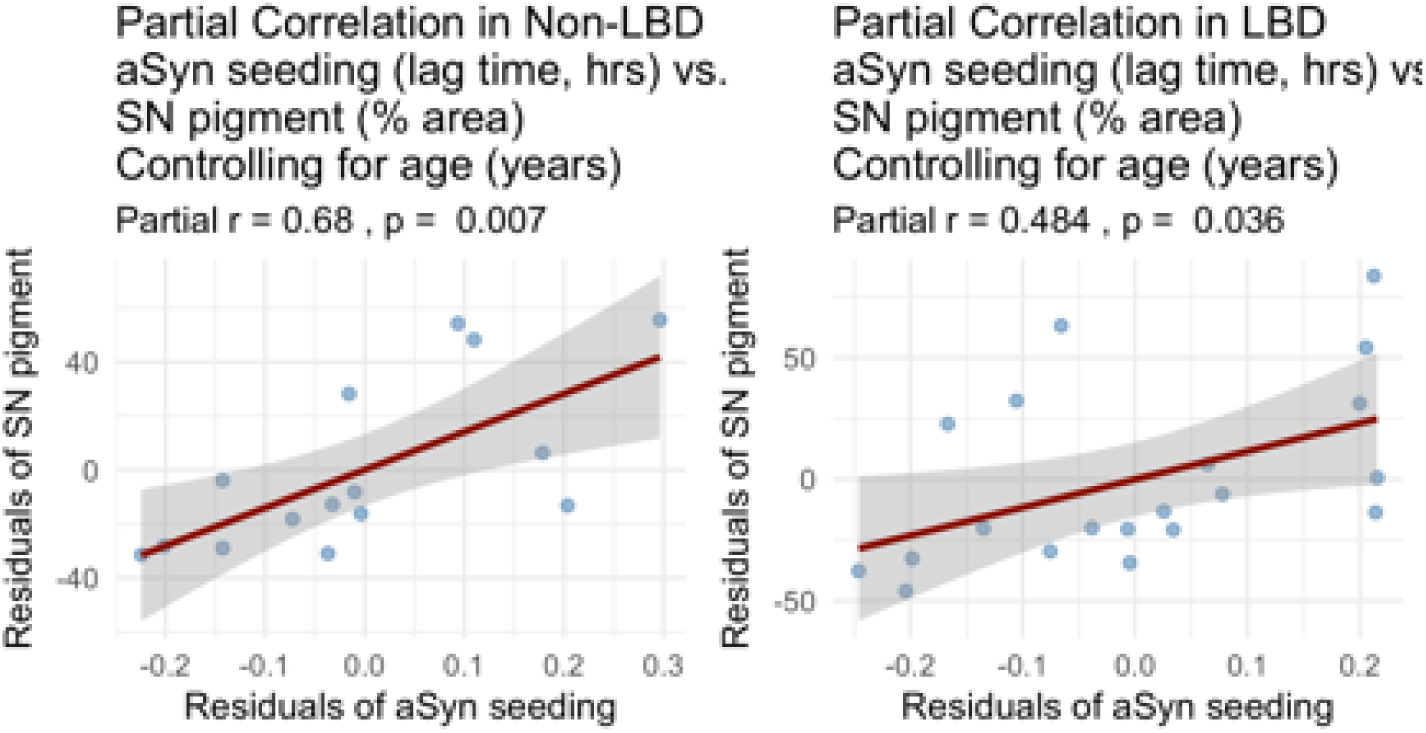
Adjustment for age using partial correlations when comparing seeded aggregation in the gut to pigment loss in the substantia nigra. Titles show partial r and p value, and graphs show residuals with linear regression (red line) for α-synuclein versus nigral pigment for both the non-LBD (left column) and LBD (right column) groups.

**sFigure 7:**
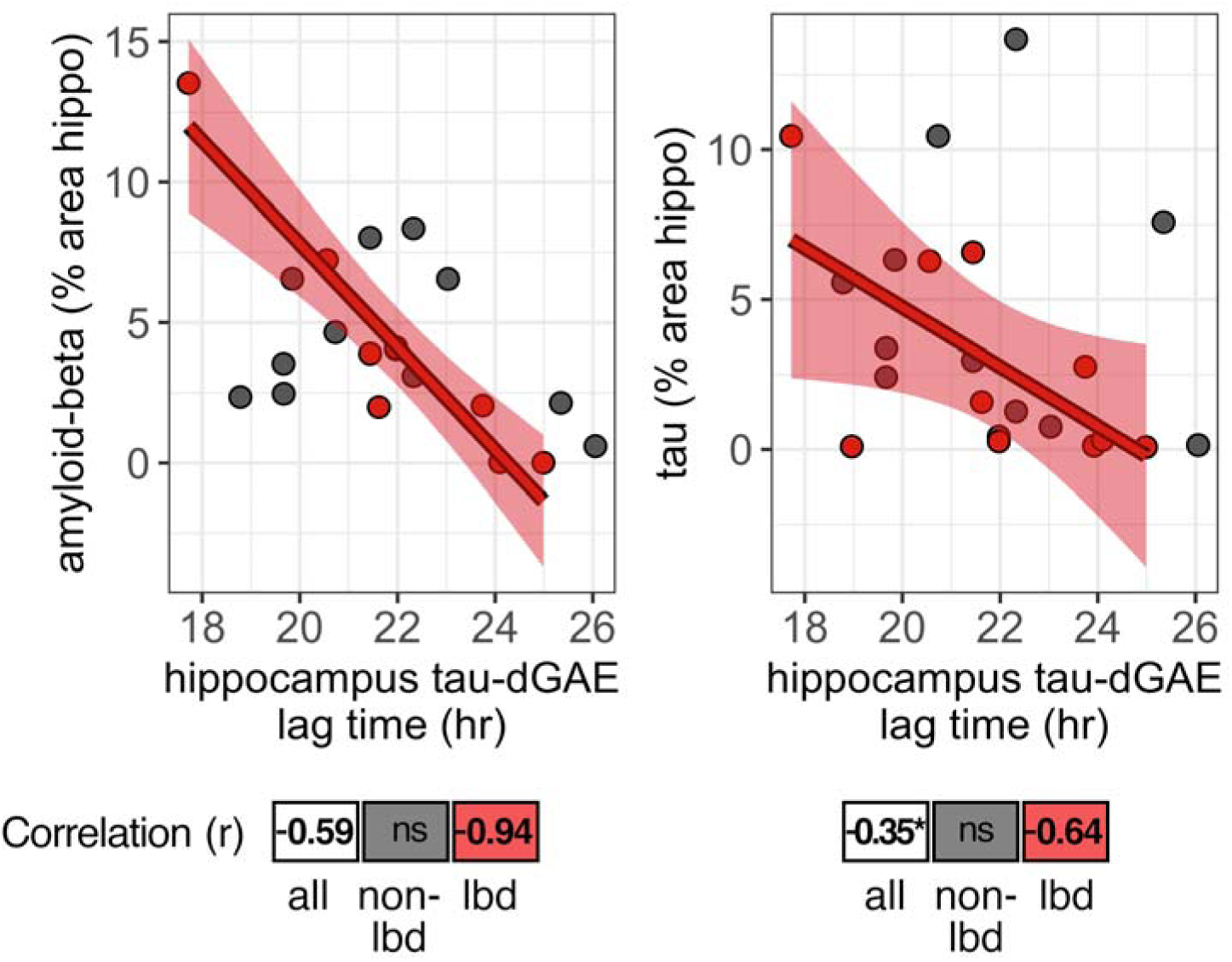
Correlation of tau-dGAE aggregation lag times in hippocampus versus quantification of immunostained amyloid-β and tau in hippocampus. Linear regression (lines, confidence intervals). Pearson r values (p < 0.05), ns = not significant. * p = 0.10

**sFigure 8:**
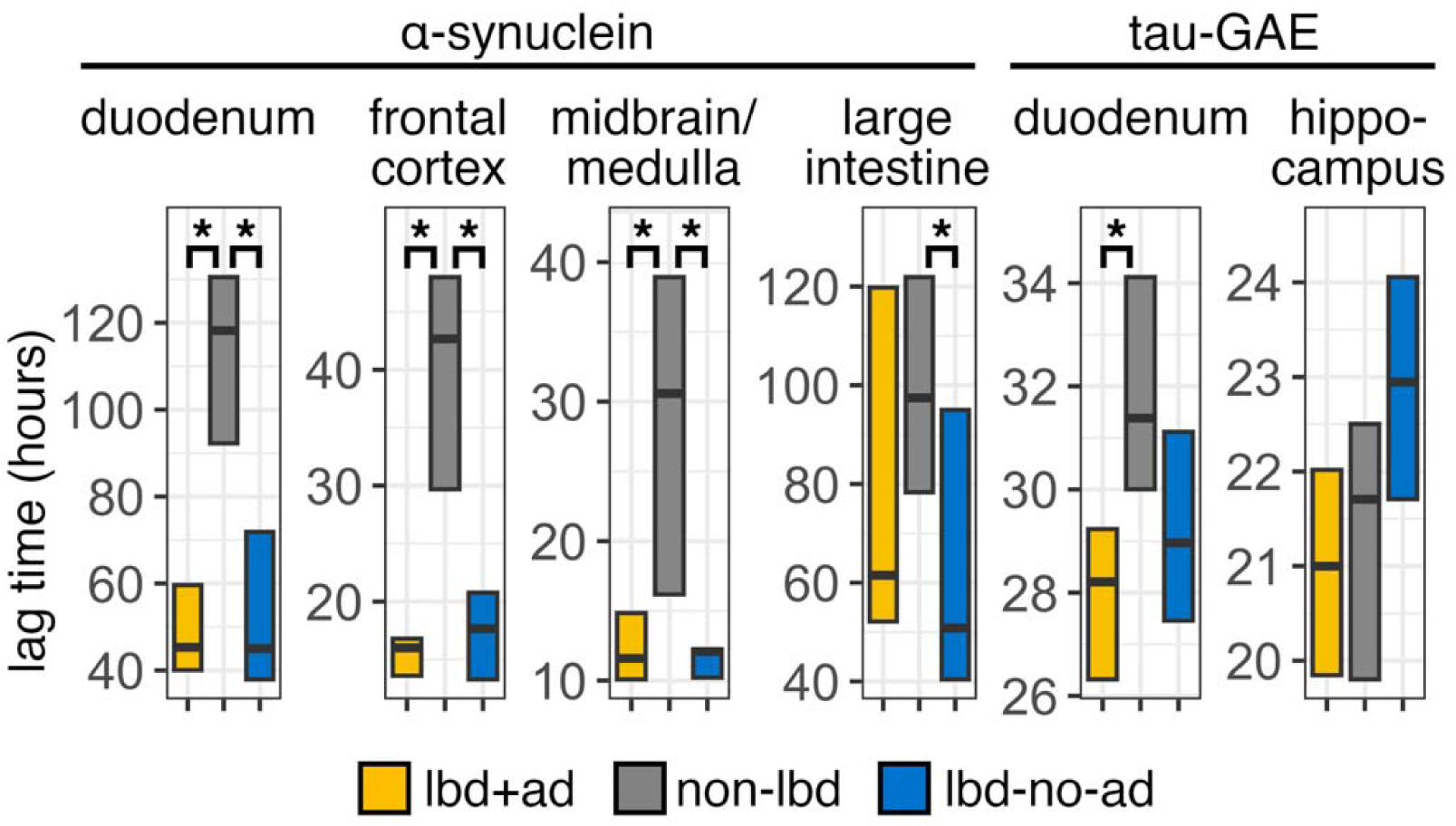
Seeded aggregation in the gastrointestinal tract and brain for subtypes of LBD with and without AD co-pathology. Boxplots show lag times for seeded aggregation of α-synuclein and tau-dGAE for participants with LBD and Alzheimer’s pathology, LBD+AD (yellow), controls without LBD (gray), and LBD without Alzheimer’s pathology, LBD-NO-AD (blue). * p < 0.05, ANOVA, Tukey post hoc test. Direct comparison of lag times in LBD+AD versus LBD-NO-AD is not significant.

## Notes

### Competing Interest Statement

Authors PK, KP, and AL are employees of StressMarq Biosciences Inc. PK, KP, and AL designed, synthesized, and provided aSyn and tau-dGAE monomers for the current study. All other authors declare no competing interests.

### Author Declarations

Institutional Review Board of Oregon Health and Science University (OHSU) gave ethical approval for this work (protocol number 1623).

### Funding

- VA Biomedical Laboratory Research and Development Career Development Award: 1 IK2 BX005760-01A1
- VA military exposures research program: I01 BX006155

